# A novel method for predicting Lp(a) levels from routine outpatient genomic testing identifies those at risk of cardiovascular disease across a diverse cohort

**DOI:** 10.1101/2024.11.01.24316526

**Authors:** Natalie Telis, Hang Dai, Ashley Waring, David Kann, Dana Wyman, Simon White, Basil Khuder, Francisco Tanudjaja, Alexandre Bolze, Matthew E. Levy, Cassie Hajek, Lisa M. McEwen, Douglas Stoller, Christopher N. Chapman, C. Anwar A. Chahal, Daniel P. Judge, Douglas A. Olson, Joseph J. Grzymski, Nicole L. Washington, William Lee, Elizabeth T. Cirulli, Shishi Luo, Kelly Schiabor Barrett

## Abstract

**Background:** Lipoprotein(a) (Lp(a)) levels are a largely genetically determined and often an unmeasured predictor of future Atherosclerotic Cardiovascular Disease (ASCVD). With the increased use of exome sequencing in the clinical setting, there is opportunity to identify patients who have a high chance of having elevated Lp(a) and are therefore at risk of ASCVD. However, accurate genetic predictors of Lp(a) are challenging to design. In addition to single nucleotide variants (SNVs), which are often summarized as a combined genetic risk score, Lp(a) levels are significantly impacted by copy number variation in repeats of the kringle IV subtype 2 domain (KIV-2), which are challenging to quantify. KIV-2 copy numbers are highly variable across populations, and understanding their impact on Lp(a) levels is important to creating an equitable and reliable genetic predictor of Lp(a)-driven cardiovascular risk for all individuals.

**Methods:** We develop a novel method to quantify individuals’ total number of KIV-2 repeats from exome data, validate this quantification against measured Lp(a) levels, and then use this method, combined with a SNV-based genetic risk score, to genotype an entire all-comers cohort of individuals from health systems across the United States (Helix Research Network; N = 76,147) for an estimated Lp(a) level.

**Results:** Our combined genotyping strategy improved prediction of those with clinically-elevated Lp(a) measurements across the genetically diverse cohort, especially for individuals not genetically similar to European reference populations, where GRS-based estimates fall short (r^2^= 0.04 for GRS, r^2^ = 0.34 KIV2+GRS in non-European). Importantly, high combined genetic risk of high Lp(a) genotypes are significantly associated with earlier onset and increased incidence in ASCVD, compared to average and low combined genetic risk genotypes in a retrospective analysis of atherosclerotic diagnoses derived from electronic health records (EHRs). This holds in the cohort at large (CAD HRs=1.29, 1.58), in the European subcohort (HRs=1.30,1.61) as well as at trending levels of significance in individuals not genetically similar to Europeans (HRs=1.22,1.31). In addition, high combined genetic risk for high Lp(a) genotypes are at least 2-fold enriched amongst individuals with ASCVD diagnosis despite a lack of EHR-based evidence of traditional risk factors for cardiovascular disease.

**Conclusions:** Our study demonstrates that genetically predicted Lp(a) levels, incorporating both SNV and our novel KIV-2 repeat estimate, may be a practical method to predict clinically elevated Lp(a). Supporting this, individuals with high combined genetic risk for high Lp(a) have an increased risk for ASCVD, as evidenced across data from seven US-based health systems.

## Introduction

Lipoprotein(a), or Lp(a), is an atherogenic lipoprotein particle comprised of two core components: low-density lipoprotein (LDL) and apolipoprotein(a), the latter of which is genetically encoded by the *LPA* gene (Marcovina and Koschinsky 1998). Circulating Lp(a) levels are predominantly determined by genetics, with the majority of variability in these levels linked to variations at the *LPA* locus (Utermann *et al*. 1987; Kraft *et al*. 1996; Clarke *et al*. 2009; Schmidt *et al*. 2016; Tsimikas and Marcovina 2022). Lp(a) levels can vary up to a hundred-fold among individuals, with different population subgroups exhibiting shifted distributions and medians(Varvel *et al*. 2016; Tsimikas 2017; Tsimikas and Marcovina 2022; Dudum *et al*. 2024).

Elevated Lp(a) levels—typically cited as roughly 120 nmol/L (50 mg/dL) or above—are present in 20% of the population and have been identified as a predictor of Atherosclerotic Cardiovascular Disease (ASCVD), independently of established ASCVD risk factors like LDL (Kamstrup *et al*. 2009; Emerging Risk Factors Collaboration *et al*. 2009; Nordestgaard *et al*. 2010; Koschinsky *et al*. 2024). While some studies suggest different clinical risk level thresholds for primary and secondary prevention of ASCVD may be warranted, there is consensus that higher levels of Lp(a) are associated with increased ASCVD risk at the population level and across ethnicities, gender, baseline ASCVD status, and subpopulations such as those with Type 2 Diabetes (T2D) (Erqou *et al*. 2010; Varvel *et al*. 2016; Patel *et al*. 2021; Berman *et al*. 2024; Wong *et al*. 2024). Aligned with this evidence, there are also emerging professional guidelines that support incorporating Lp(a) screening strategies. Both universal, one-time Lp(a) screening for adults or screening of those at intermediate or high risk via established CAD risk assessments have been recommended (Grundy *et al*. 2019; Cegla *et al*. 2019; Tsimikas and Stroes 2020; Koschinsky *et al*. 2024).

Lp(a) is not typically measured in the course of care, and accurate measurement of Lp(a) levels across the spectrum of Lp(a) isoform sizes is difficult and a known limitation of current laboratory tests (Wilson *et al*. 2019). Given the high heritability of Lp(a) levels, genetic estimation of Lp(a) has the potential to be a practical alternative to more traditional lab measurement, especially when screening populations (Reyes-Soffer *et al*. 2022; Trinder *et al*. 2022). Both an SNV-based genetic risk score (GRS) and KIV-2 copy number have independently been used to genetically estimate clinically-relevant Lp(a) levels (Kamstrup *et al*. 2009; Burgess *et al*. 2018; Trinder *et al*. 2020). While both axes of variation contribute to circulating Lp(a) levels, due to genotyping and sequencing prerequisites for each variation type, the SNV-based GRS and KIV-2 copy number variation have seldom been studied together, particularly in large cohorts(Mack *et al*. 2017; Zekavat *et al*. 2018).

It has been shown that the SNV-based GRS associates with ASCVD for individuals who self-reported their ethnicity to be European; however, this same GRS was not evaluated in those who self-reported other ethnicities, limiting the clinical utility of such estimations for Lp(a)-related disease risk (Trinder *et al*. 2020). Recent research datasets, comprising whole genome or targeted *LPA* locus sequencing data and measured Lp(a) levels, illustrate the potential benefits of incorporating both SNV and KIV-2 copy number variations to genetically estimate Lp(a)(Zekavat *et al*. 2018; Coassin and Kronenberg 2022). The improvements in *LPA* genetic estimations are particularly noticeable in participants with self-reported non-European ancestries, where smaller KIV-2 copy number alleles, which contribute to clinically significant Lp(a) elevations, are more prevalent (Mehta *et al*. 2022; Tsimikas and Marcovina 2022).

We genotype a large, diverse clinicogenomic cohort derived from seven health systems across the United States–the Helix Research Network (HRN)--for both *LPA* GRS and estimated total KIV-2 CNV count. We show that, in the subset of individuals with measured Lp(a) values in their medical records, using the combination of these genetic elements to predict those with Lp(a) levels in the clinically-elevated range is much more accurate than using either element alone. To test clinical utility of these genetic-based Lp(a) predictions across ancestries, we then use GRS and the estimated number of KIV-2 repeats to establish low, average and high combined genetic risk Lp(a) groups across the cohort and associate these risk groups to cardiovascular phenotypes with established connections to Lp(a) levels (Clarke *et al*. 2009; Klarin *et al*. 2019; Kronenberg *et al*. 2022).

## Methods

### Subjects and genetic data

Our study comprised 76,147 genetic samples from participants in the Helix Research Network (HRN) that were sequenced and analyzed at Helix using the Exome+^®^ assay as previously described, recruited from the Healthy Nevada Project (n=40,430); myGenetics (n=23,113); and In Our DNA SC (n=12,604) (see Supplementary Table 1) (Cirulli *et al*. 2020). The HRN studies were reviewed and approved by their applicable Institutional Review Boards (IRB, projects 956068-12 and 21143). All participants gave their informed, written consent prior to participation. All data used for research were deidentified. Demographic distributions of these individuals may be found in Supplementary Tables 1-3.

### Phenotypes

HRN phenotypes were processed from Epic/Clarity Electronic Health Records (EHR) data as previously described and updated as of January 2024 (Cirulli *et al*. 2020). International Classification of Diseases, Ninth and Tenth Revision, US Clinical Modification (ICD-9-CM and ICD-10-CM) codes and associated dates were collected from available diagnosis tables (from problem lists, medical histories, admissions data, surgical case data, account data, and invoices). All datasets were transformed into the OMOP Common Data Model v5.4. Therefore, diagnosis codes were mapped to standard SNOMED terms and represented by concept ids. To be comprehensive in our cohort definition, we used both the source concept id and the standard concept id to extract relevant diagnoses.

Coronary artery disease (CAD), peripheral artery disease (PAD), aortic stenosis (AS), Ischemic Stroke (IS), and myocardial infarction (MI) were defined through review of each respective disease diagnosis in the phecode X map(Shuey *et al*. 2023), physician input, as well as a literature search around clinical diagnosis codes (ICD-10, ICD-10-CM, ICD-9-CM, SNOMED) given the different natures of the source datasets. We identified the first record of each included ICD-10, ICD-10-CM, ICD9-CM or SNOMED term for each disease phenotype to represent the first occurrence of disease for the associated individual. A complete list of diagnosis codes included in each phenotype definition may be found in Appendix Table 1.

### Clinical measurements

HDL, LDL and Lp(a) measurements were identified in the HRN as any measurement matching a set of codes to exhaustively capture blood assays encoded in different units (see Appendix Table 2). Each measurement was transformed into a unified set of units. When measured in mg/dL, Lp(a) was converted to nmol/L using a conversion factor of 2.15 as established in the literature (see for example (Khera *et al*. 2014).

### Statistical analyses

Statistical analyses were run using the statsmodel package in Python 3.7.3. For binary variables, logistic regression or Firth logistic regression was used; for quantitative variables, linear regression was used after rank-based inverse normal transformation. For time to event analyses, the lifelines package was used, using age of first diagnosis received anywhere in their medical record as an end point. Cox proportional hazards were adjusted for sex, age, population group, and BMI, a common cardiovascular disease risk factor available for most participants (n with BMI= 101,986 [94%]). Linear models were adjusted for the exome assay version and KIV-2 copy number estimate (CNE) was normalized for each exome assay subset (see Supplementary Figure 3).

### Genetic similarity

Genetic similarity was assigned using ADMIXTURE with the 1000 Genomes Project (1KGP) population set as a reference (Alexander *et al*. 2009; 1000 Genomes Project Consortium *et al*. 2015). To facilitate comparisons with previous studies, and to report results in populations previously underrepresented cohorts in genetics research, we divided our cohort into European (EUR) and non-European (non-EUR) groups as follows. For each participant, we used exome-wide variant calls to calculate Admixture coefficients for five continental supergroups -- AFR, AMR, EAS, EUR, SAS -- derived from data unifying 26 separate populations in the Phase 3 data of the 1000 Genomes project. Our EUR cohort includes only individuals with EUR > 0.85 (a majority of their genetic similarity assigned to the European populations; and non-EUR < 0.1), and non-EUR describes individuals with a majority of their ancestry assigned to any of the other population groups (‘AFR’, ‘AMR’, ‘EAS’, ‘SAS’). A full breakdown of population assignment and demographics for the full cohort, as well as the subset with Lp(a) measurements, can be found in Supplementary Table 1.

This approach follows guidelines on the use of population descriptors in genetics research, where the EUR group represents individuals genetically similar to the EUR reference group from the 1000 Genomes project and the non-EUR group represents individuals that are not genetically similar to the EUR reference group.

### Genetic measures associated with Lp(a)

We calculate a genetic risk score (GRS) to predict Lp(a) levels, using genetic variants and corresponding effect estimates from (Trinder *et al*. 2020), PGS Catalog # PGP000127).

### KIV-2 normalized coverage based copy number quantification

Exact KIV-2 copy numbers for both alleles are challenging to determine from short-read whole exome sequencing data. However, a quantification of the total KIV-2 copy number difference among individuals sequenced by the same assay can still be used as a numerical variable to stratify variation in KIV-2 copy number for each individual. Here we developed a method to quantify such a difference.

For any individual, an *LPA* allele is expected to have one copy for each of the KIV-1 and KIV-3 to KIV-10 domains, but an unknown number of copies of the KIV-2 domain. On reference genome hg38, the *LPA* gene has six copies of KIV-2 domain with identical or slightly different sequences, so we masked the second to sixth KIV-2 domains; we also masked the second exon of the KIV-1 domain and the first exon of the KIV-3 domain due to their similarity to KIV-2. Reads extracted from *LPA* are re-aligned to the masked chr6 reference sequence, and the coverage is counted for unmasked exons of KIV domains.

The total copy number of KIV-2, estimated through KIV-2 exon 1, is quantified as the following normalized coverage of KIV-2 exon 1:

Use **KIV-i-1** to denote the coverage of KIV-i domain exon 1,

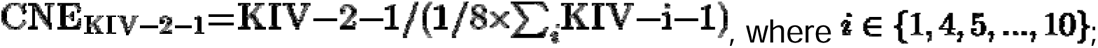

Similarly, that estimated through KIV-2 exon 2 is quantified as:

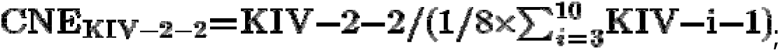

and the final total copy number of KIV-2 is quantified as:

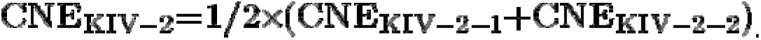

Afterwards, this copy number of KIV-2 is normalized by exome assay version to account for variability in region coverage, forming the final KIV-2 CNE.

### Assigning genetic risk of clinically elevated Lp(a)

In the distribution of individuals with empirically measured Lp(a), we set out to determine the GRS as well as KIV-2 CNE genetic profiles of those at the extreme ends of the spectrum, which represent clinically relevant ranges. We define combined cutoffs for the GRS as well as KIV-2 CNE to identify individuals with a high, medium, and low genetic risk of clinically elevated Lp(a). For the GRS, for high risk we choose a threshold of 120 nmol/L, as the GRS effect sizes are measured in units of nmol/L, to reflect the clinically significant level. For KIV-2 CNE, as it is normalized per version of our exome (see above), we choose a threshold of +/-0.8 (KIV-2 CNE above or below 0.8 standard deviations from the mean) to match the proportion of individuals with a measured Lp(a) above 120 nmol/L. High risk is defined as either a GRS > 120 nmol/L or a KIV-2 CNE below 0.8 standard deviations from the mean. Low risk is defined as either a GRS <10nmol/L or a KIV-2 CNE above 0.8 standard deviations from the mean. All other genotype combinations are considered average risk.

To extrapolate to the general population, a slightly different approach is needed, as the distribution of Lp(a) measures taken due to clinical care is ascertained and therefore lopsided upward relative to the expected range. Nonetheless, we continue to focus on the extremes of the genetic distributions on either end. We continue using the GRS thresholds of >120 nmol/L and <10 nmol/L for high and low risk and match a KIV-2 CNE cutoff to reflect this proportion in the general population, resulting in a cutoff of +/- 1.5 SD, somewhat more conservative than that used in analyses of the subcohort with Lp(a) measures. This ensures individuals at high and low ends of genetic risk of high Lp(a) are included at a roughly similar rate and that GRS and KIV-2 CNE are both contributing to the combined genotypes. These high and low combined genotypes reflect our best understanding of clinical risk of ASCVD, but we also evaluated a threshold corresponding to 150nmol/L, to represent a significant clinical endpoint (for example, see (Yeang *et al*. 2022). The demographic distribution of the resulting risk groups calculated using either a GRS cutoff of 120 nmol/L and 150nmol/L determining the KIV-2 CNE distribution cutoffs can be found in Supplementary Table 3.

## Results

### Lp(a) is seldom measured in clinical practice

Within an all-comers cohort recruited from three health systems in the Helix Research Network (HRN, n=76,147), we identify only 1584 (2.0%) individuals with 1777 clinically ascertained Lp(a) measurements (for a full breakdown of cohort demographics, see Supplementary Table 1). In contrast, 99% of individuals across this cohort have an LDL measurement. As these Lp(a) measurements were taken during routine care in response to a clinical observation, we expect them to be non-representative of the total population (Varvel *et al*. 2016; Dudum *et al*. 2024). Indeed, the mean Lp(a) measured in the cohort is 50 nmol/L, slightly below the upper threshold for normal (<75 nmol/L) and higher than the expected population mean (28-45 nmol/L) (Langsted *et al*. 2014; Patel *et al*. 2021). This cohort also has a slightly elevated LDL when measured (mean 101.4 mg/dL; for full comparison see Supplementary Figure 4).

### SNV and KIV-2-based genetic predictor performance in those with Lp(a) levels

To facilitate comparisons with previous studies, and to report results in populations previously underrepresented cohorts in genetics research, we divided our cohort into European (EUR) and non-European (non-EUR) groups (see Methods). As expected, a previously-validated genetic risk score (GRS), which outputs a genetically predicted Lp(a) level based on SNVs in *LPA,* is associated with Lp(a) levels in EUR individuals in our cohort (n=1346) with an r^2^ of 0.4 (see Supplemental Figure 1). However, the r^2^ in non-EUR individuals (n=238) is 0.04, indicating little predictive value of the GRS for Lp(a) levels in non-EURs. In parallel, KIV-2 copy number estimation (CNE) is anticorrelated with Lp(a) levels in both subsets, with an r^2^ of 0.32 and 0.29 for EUR and non-EUR cohorts, respectively (see Supplemental Figure 2). This negative correlation is expected as a lower KIV-2 CNE implies a smaller total molecule size, which eases export of the molecule, increasing measured blood levels of Lp(a) (Reyes-Soffer *et al*. 2022). Accordingly, a linear model combining both genetic predictors to genetically estimate Lp(a) improves performance, with an r^2^ of 0.49 for EUR individuals (p<2.2e-16 for both variables) and 0.34 for non-EURs (p<2.2e-16 for KIV-2 CNE; p=5.5e-6 for GRS) in the dataset; this model performs better than either alone.

As a performance metric, the correlation coefficient measures the ability to predict the specific Lp(a) values in a patient population. However, the key clinical outcome for population-wide genetic screening is whether an individual is expected to have an Lp(a) level high enough to be associated with excess risk of future cardiovascular morbidity (Grundy *et al*. 2019; Kronenberg *et al*. 2022). Accordingly, we calculate an additional performance metric to determine if KIV-2 CNE and GRS genotype groups can be established for predicting clinically relevant Lp(a) elevations (>120 nmol/L). We identify individuals with a high GRS or a low KIV-2 CNE by extrapolating from the distribution for individuals with a clinically elevated Lp(a) (see Methods). In the subset of individuals with low KIV-2 CNE (approximately the lower 20% of the distribution), 70% of these individuals have a clinically elevated Lp(a), compared to only 17% of people without this genetic risk. Likewise for individuals with high GRS (above 120 nmol/L predicted), 80% have a clinically elevated Lp(a), compared to only 16% without (Figure 1, Chi squared p-value < 2.2e-16). The GRS performance differs significantly by population; 88% of EUR individuals with a high GRS have an elevated Lp(a), compared to only 43% of non-EUR individuals. When unifying across the presence of either genetic risk, we find 89% of individuals with either a high GRS or a low KIV-2 CNE have a clinically elevated Lp(a), compared to only 9.8% of individuals with neither genetic risk predictor. This combined model performed similarly well for individuals of both the EUR and non-EUR cohorts, in stark contrast to the ancestry-biased performance of the GRS.

**Figure 1:**
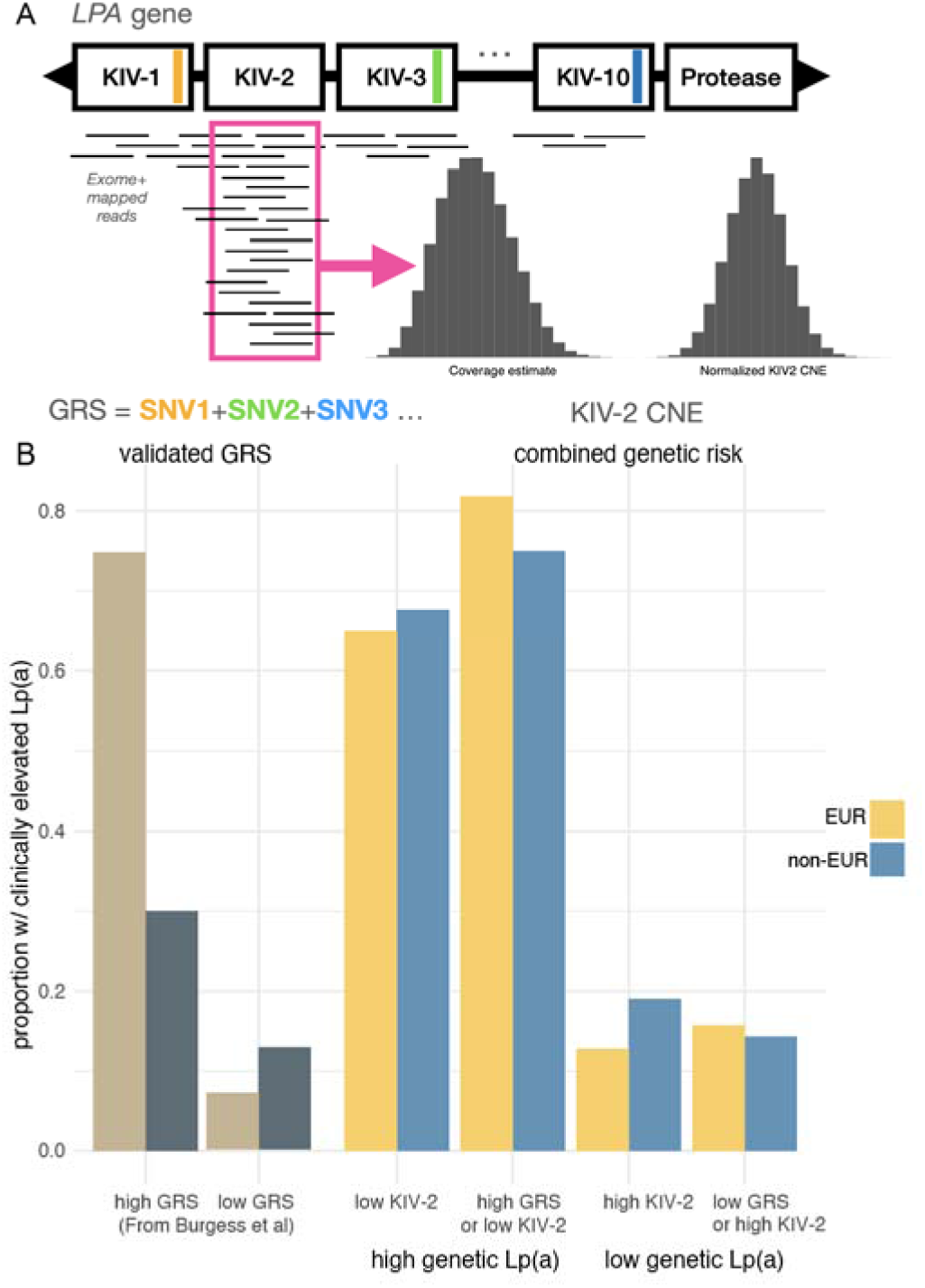
GRS and KIV-2 each contribute independently to predicting Lp(a) levels. A) Schematic representation of bioinformatic process for KIV-2 copy number estimate (CNE) and calculation of th *LPA* GRS score. B) Proportion of individuals in each category with an Lp(a) > 120nmol/L, divided by membership i either EUR (‘European’) or non-EUR (‘Non Eur’) cohort. From left to right, ‘high GRS’ is a GRS > 120 nmol/L (as the GRS is in units of predicted Lp(a)); ‘low KIV-2’ denotes a normalized KIV-2 less than 0.8 standard deviations from the mean; and ‘high GRS or low KIV-2’ indicates that a patient was included for either reason. Although some individuals with high GRS may have a non-risk KIV-2, and vice versa, ‘low PRS or high KIV-2’ are individuals with neither genetic risk factor.

### Combined genetic risk for predicting clinically-relevant Lp(a) levels

Guided by this data, we developed a population-level, ancestry-agnostic genotyping approach for categorizing genetic Lp(a) risk leveraging both our KIV-2 CNE and the *LPA* GRS. Specifically, we assign individuals to have a combined genetic risk for a high Lp(a) (“high combined genetic risk”) if they have either a GRS >120 nmol/L or a KIV-2 CNE below 1.5 standard deviations from the mean (n=14,355, or 18% of the participants). The remaining individuals are considered to have a low combined genetic risk for a high Lp(a) (“low combined genetic risk”) if they have a GRS < 10 nmol/L or a KIV-2 CNE above 1.5 standard deviations from the mean (n=8,659). The remaining individuals harbor genetic profiles indicative of average Lp(a) levels (n=53,133). Notably, the average LDL for individuals with recorded lab values is consistent between these genetic groups, ranging between 104-107 (Supplementary Figure 4).

### Combined genetic risk, predicting clinically-elevated Lp(a), is associated with increased ASCVD diagnoses in both EUR and non-EUR individuals

Next, we investigate association of our combined genetic risk groups with cardiovascular disease diagnoses associated with clinically-elevated Lp(a) including coronary artery disease (CAD), peripheral artery disease (PAD), myocardial infarction (MI), aortic stenosis (AS), ischemic stroke (IS), as well as the five-point Major Adverse Cardiovascular Event (MACE) index. For each phenotype, we compare the rate and age of onset of these diagnoses between the high and average and high and low combined genetic risk groups using a Cox regression controlling for sex, age, population group, and BMI to calculate a proportional hazard ratio (HR) for each comparison. We find significant proportional hazards for CAD, PAD, MI, AS and MACE, with an increase in the magnitude of these differences when comparing the high combined genetic risk group to the average combined genetic risk group (e.g. CAD HR=1.29 [1.22-1.38], p= 2.01e-16; PAD HR=1.34 [1.08-1.66], p = 0.0015) and low combined genetic risk group (CAD HR= 1.58[1.43-1.75], p=4.89 e-18; PAD HR= 2.05 [1.39-3.03], p=6.8e-5) (See Figure 2 and Appendix 2 for all pairwise combinations). IS was only moderately enriched in the high combined genetic risk group compared to the low combined genetic risk group (HR = 1.19[1.02-1.41], p = 0.03).

**Figure 2.**
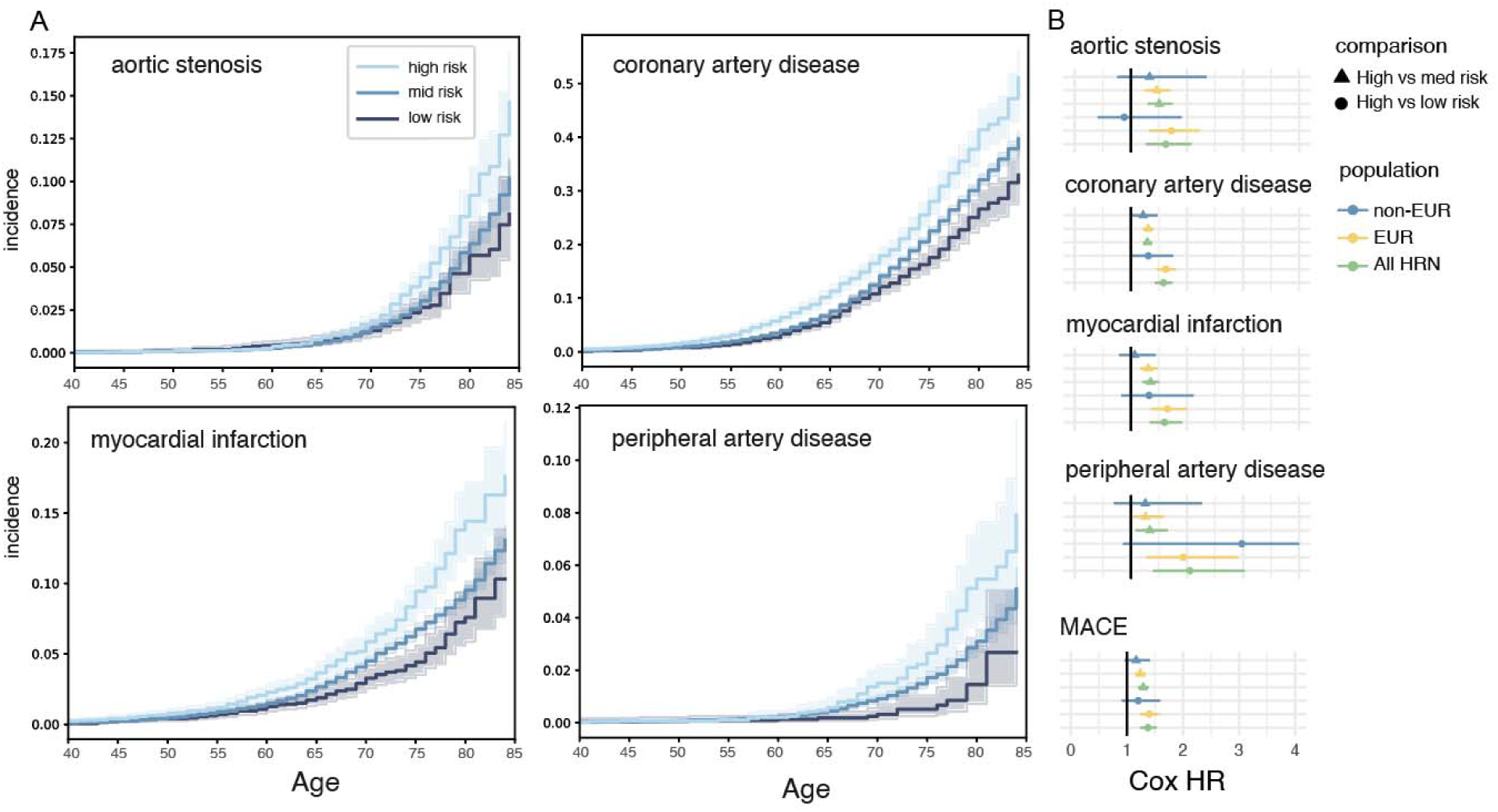
Elevated combined genetic risk of a high Lp(a) is associated with increased phenotype incidence after adjusting for age, sex, BMI and population of origin. A) the cumulative fraction of individuals with coronary artery disease, divided by their Lp(a) combined genetic risk group (high, average, or low). B) The Cox proportional hazar ratio and 95% confidence intervals for high versus average or high versus low combined genetic risk of high Lp(a) groups across statistical significant phenotypes in the unified cohort, as well as these estimations in subcohorts separated by genetic similarity (EUR or non-EUR cohorts).

Next, we re-test for association in EUR (n=61,406) and non-EUR (n=14,741) cohorts separately. As expected, given sample size enrichment (81% of cohort) and established performance of GRS against similar outcomes, all comparisons remain significant in the EUR subset(Figure 3B, Appendix 2). Despite the substantially smaller sample sizes of the high (n=3,349) and low (n=1,798) combined genetic risk groups for non-EUR cohort, we observe similar stepwise increases in hazard ratio confidence intervals for the suite of phenotypes with significant HR differences in the cohort at large (Figure 3B, Appendix 2). In particular, differences in diagnoses trends for CAD between the high and average combined genetic risk groups and high and low combined genetic risk groups trend towards significance (HR=1.22[1.0-1.48], p=0.04; HR=1.31[0.98-1.76], p=0.06). The exception was AS in the high vs. the low combined genetic risk group for non-EUR, where low power was an issue (36% power to detect the effect size seen in the EUR group in the non-EUR group). Nonetheless, without the use of a clinical Lp(a) measurement, these genetically predicted Lp(a) risk groups identify individuals with an increased incidence of future cardiovascular morbidity.

**Figure 3:**
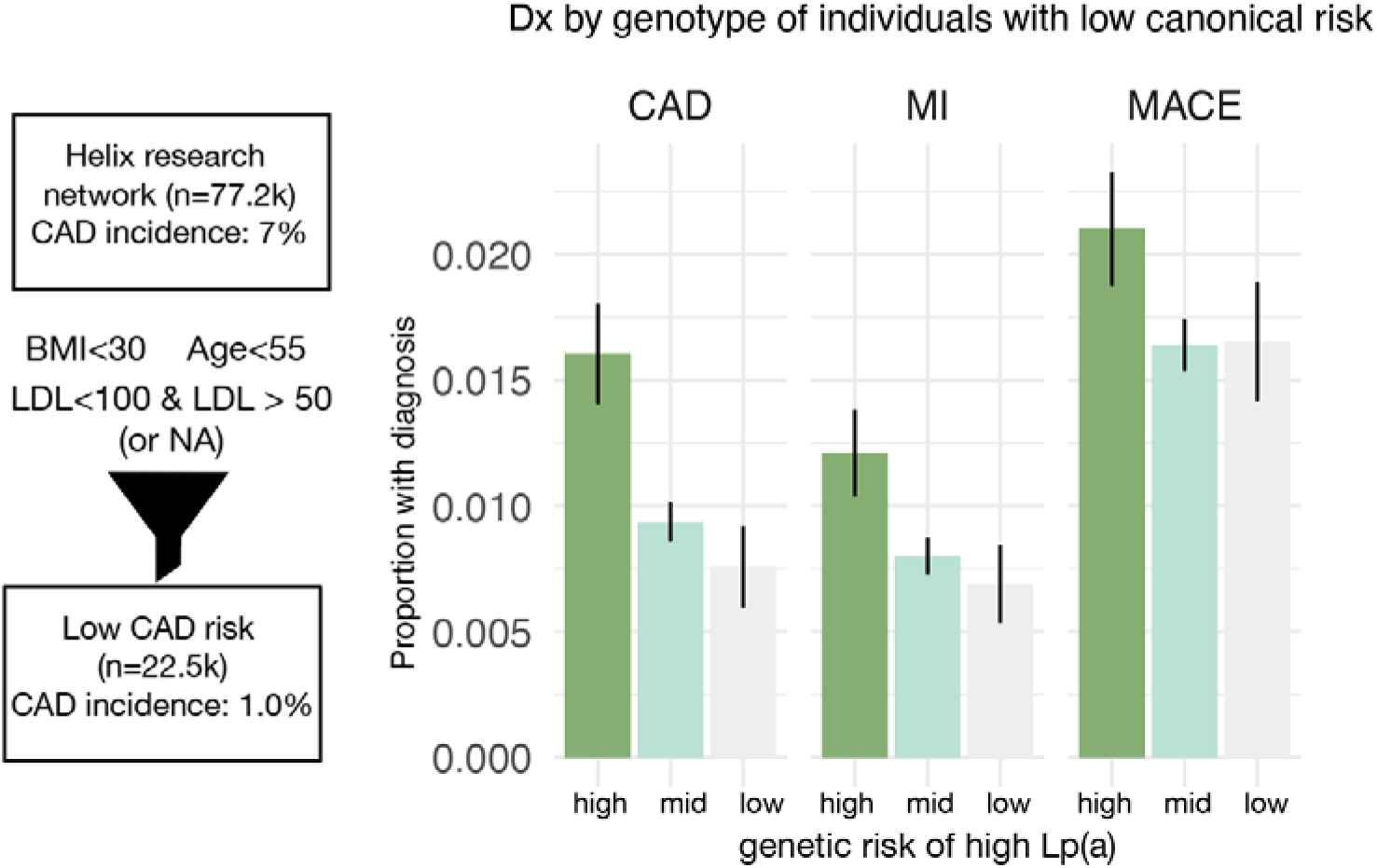
Proportion of individuals at low canonical risk with disease diagnoses. The proportion of individuals in eac Lp(a) genetic risk group receiving a diagnosis. These individuals are all restricted to age <55, BMI < 30, and either n LDL measurement or a low measurement, to include only individuals with no canonical risk factors for cardiovascular disease.

### Predicted Lp(a) and incidence of ASCVD in individuals with otherwise low risk

In addition to increasing ASCVD risk independently of other risk factors like LDL, Lp(a) is also suspected to contribute to the incidence of premature or otherwise unexpected ASCVD (Verbeek *et al*. 2018). To measure the latent contribution of a high combined genetic risk of high Lp(a) to unexpected ASCVD, we subsetted our cohort to individuals with a lower risk of cardiovascular disease using key risk factors available from EHR records. We included individuals with either no LDL measurement or a low clinically-ascertained measurement (<100 mg/dL, but >50mg/dL, to exclude people with other possible lipid disorders), low age (<55), and most recent BMI < 30 (Zhou *et al*. 2021; Wong *et al*. 2022). As expected, this subset has a substantially lowered incidence of CAD (Figure 3). Next, we compared the percentage of individuals in each combined genetic risk category with an early ASCVD diagnosis (age <55). The mean age in each risk category in this analysis was similar (41.4, 41.5, and 41.4 years across high, medium, and low risk groups; see Supplemental Figure 5) so comparing incidence across groups is not biased by age of onset or followup time. We find that 1.2-1.6% of individuals with a high combined genetic risk of a high Lp(a) in this low traditional risk cohort harbor a diagnosis of CAD or MI, compared to 0.6-0.7% of low traditional risk individuals with a low combined genetic risk of high Lp(a) (Figure 3).

Another way to summarize this contribution to excess risk is to calculate the proportion of individuals in high combined genetic risk of high Lp(a) receiving premature diagnoses compared to the overall representation of the group in the population. Using CAD as an example, we find that while individuals with a high combined genetic risk of high Lp(a) are 17% of the low risk population, they comprise 27% of low canonical risk individuals who are nonetheless diagnosed with CAD (binomial test, *p*-value = 0.014). Across ASCVD diagnoses in this low risk population, there is 2-fold enrichment in those with high combined genetic risk of high Lp(a) compared to those with low combined genetic risk of Lp(a) (Figure 3). While these enrichments may not account for the majority of cardiovascular morbidity, elevated Lp(a) represents an unobserved determinant of risk, and may explain nearly a third of individuals diagnosed with disease who lack other canonical risk factors.

## Discussion

Genetic prediction of Lp(a) levels presents significant challenges as well as significant potential for precision medicine and prevention of future risk of Atherosclerotic Cardiovascular Disease (ASCVD). In this work, we develop a practical method to estimate the number of copies of KIV-2 (KIV-2 CNE) from exome sequencing in the Helix Research Network, an all-comers cohort collected from multiple health system sites across the United States. While genetic risk scores (GRS) applied in the clinic are known to perform poorly in understudied and underrepresented genetic groups, KIV-2 CNE is a population-agnostic genetic risk predictor (Privé *et al*. 2022; Kullo *et al*. 2022). KIV-2 CNE performed well to predict Lp(a) in both the Europrean (EUR) and non-European (non-EUR) subsets of our cohort with Lp(a) measurements (Figure 1). Independent of genetic similarity group, individuals with a low KIV-2 CNE are 5-fold more likely to present with elevated Lp(a) levels (80% vs 16%, Chi squared p-value < 2.2e-16). This demonstrates the opportunity to develop a scalable genetic risk predictor and with equitable performance, which we test across a diverse health system cohort.

We use KIV-2 CNE in combination with a GRS to develop a combined genetic risk predictor of high Lp(a) and genotype a cohort of 76,417 individuals for this genetic risk. This exome-based categorization of combined genetic risk of high Lp(a) is associated with increased incidence and earlier onset of coronary artery disease, myocardial infarction, heart failure, and other phenotypes associated with ASCVD with strong statistical significance in both the cohort at large, the EUR subset, and with trending significance and similar direction of effects in the non-EUR subcohort (Figure 2). Encouragingly, the hazard ratio estimate for our combined high genetic risk group compared to the low risk group for coronary artery disease (HR=1.6, whole cohort; p-value = 2e-19) is similar to the established hazard ratios from association studies based on actual measures of clinically-elevated Lp(a), confirming that this genetic estimate is a useful clinical instrument for predicting elevated Lp(a) levels (Finneran *et al*. 2021).

Lp(a) has been shown to predict cardiovascular morbidity risk independently of leading risk factors, such as LDL, age, and BMI, as well as in individuals who develop ASCVD without any of these canonical risk factors. Amongst patients at low canonical risk in our cohort, there is a 2-fold enrichment of individuals with a high combined genetic risk of high Lp(a) compared to low combined genetic risk individuals, and this high risk group accounts for 27% of unexpected cases of coronary artery disease (Figure 3). This suggests that our genetic measure may help identify additional individuals with elevated ASCVD risk not identified based on standard risk scoring measures.

This study does not exhaustively look at all possible genetic variation in Lp(a) with an impact on Lp(a) levels; recent work has shown that SNVs in the KIV-2 repeat itself are impactful, although likely less impactful on Lp(a) than population-specific variation in the abundance of KIV-2 (Di Maio *et al*. 2020; Schachtl-Riess *et al*. 2021; Behera *et al*. 2023). Future work incorporating SNVs, particularly in the KIV-2 region, as well as tailoring the distribution thresholds for high and low KIV-2 copy number estimates may improve the precision of these genetic estimations. One of the major limitations of studies that rely on retrospective analyses of EHR is the lack of detailed phenotyping over a longitudinal timespan and inherent selection bias in building a dataset from participants actively engaging with the healthcare system. Although EHRs are very comprehensive, many data points are not recorded with the desired frequency or detail, and coding patterns vary by both physician and health system and will be biased by the overall engagement of each participant with the health system, making it challenging to comprehensively apply known ASCVD risk estimates (D’Agostino *et al*. 2008). Despite these limitations, using EHR data for phenotyping provides an opportunity to evaluate genetic associations with health outcomes at a population level, creating evidence to support more detailed clinical follow up projects. For example, based on this retrospective analysis, we believe a prospective analysis validating this combined genetic estimation of high Lp(a) against clinically-measured Lp(a) levels more broadly across populations is warranted based on these preliminary findings.

Therapy options for patients with elevated Lp(a) are limited. Current available lipid lowering drugs do not provide targeted Lp(a) lowering. PCSK9 monoclonal antibody inhibitors modestly decrease Lp(a) levels(Burgess *et al*. 2018; O’Donoghue *et al*. 2019; Lamina *et al*. 2019; Bittner *et al*. 2020). In patients with clinical ASCVD and recurrent events, if Lp(a) is elevated, apheresis can be considered; however, this is a cumbersome and undesirable option for most patients (Leebmann *et al*. 2013; Pokrovsky *et al*. 2016). Multiple novel nucleic acid therapies that lower Lp(a) are currently being studied in phase III clinical outcomes trials (Tsimikas *et al*. 2020; O’Donoghue *et al*. 2022). Pelacarsen, an antisense oligonucleotide, and olezarsen, a small interfering RNA (siRNA), have been found to safely reduce Lp(a) by 40-80% depending on the dose and frequency of injections. The HORIZON and OCEAN(a) trials will determine if these medications reduce cardiovascular events in patients with a history of clinical ASCVD; these results are expected as early as 2025. Additionally, the ACCLAIM-Lp(a) trial will investigate lepodisiran, another siRNA that reduces Lp(a), in a phase III clinical outcomes trial for both primary and secondary prevention patients. Currently, Lp(a) measurement is used mostly for ASCVD risk stratification, but if these pharmaceutical options prove to reduce cardiovascular events, there will be an even greater interest in identifying patients with elevated Lp(a) for prevention.

Lp(a) is rarely measured as part of clinical practice: in our clinical cohort, only 1.4% of patients had a documented Lp(a) measurement taken. In contrast, every one of them has been sequenced. As sequencing becomes an established part of routine care, the impact of genetic data on treatment and preventative medicine will be determined by the efficacy and fairness of genomic predictors of future morbidity. Accordingly, we develop a predictor which performs similarly to the value of the actual lab measurement in patients where it has been ascertained, and which is highly sensitive regardless of ancestry and population of origin. This cross-population performance is crucial to the fair and equitable development of genetic medicine, and is rarely achieved in the development of polygenic predictors (Wang *et al*. 2020; Privé *et al*. 2022; Kullo *et al*. 2022; Aw *et al*. 2024). This predictor enables us to identify patients at significantly higher risk of future cardiovascular disease, which can both compound traditional risk as well as identify at risk individuals who present without canonical risk factors. In short, this genomic-based predictor for high Lp(a), which may be calculated for any patient regardless of current diagnoses, family history, or other measured risk factors, is both a fair future predictor of cardiovascular morbidity and demonstrates that 27% of unexplained coronary artery disease may be attributed to a combined genetic risk of elevated Lp(a). This work advances the evidence base supporting Lp(a) estimation as a potential equitable baseline health measure from genetic data, especially as exome sequencing for disease risk estimation and prevention becomes more common in medicine.

## Supporting information

Appendix 1 (codesets)

Appendix 2 (hazard ratios)

## Data Availability

Deidentified data may be made available upon reasonable request to the authors.

## Acknowledgments

We acknowledge the entire Helix bioinformatics and lab teams for their contributions to the production of exome sequencing pipeline as well as the research administration team for coordinating the project. We thank all of the participants of the Helix Research Network.

## Ethics Declaration

The Helix cohorts were reviewed by Salus IRB (Reliance on Salus for all sites) and approved (approval number 21143), the WCG IRB (Western Institutional Review Board, WIRB-Copernicus Group) and approved (approval number 20224919), the MUSC Institutional Review Board for Human Research and approved (approval number Pro00129083), and the University of Nevada, Reno Institutional Review Board and approved (approval number 7701703417). All participants gave their informed, written consent before participation. All data used for research were de-identified.

