## Supplementary tables and figures for "A novel method for predicting Lp(a) levels from routine outpatient genomic testing identifies those at risk of cardiovascular disease across a diverse cohort"

Figure 1. Scatterplot of Lp(a) vs GRS (calculated as in Trinder et al)

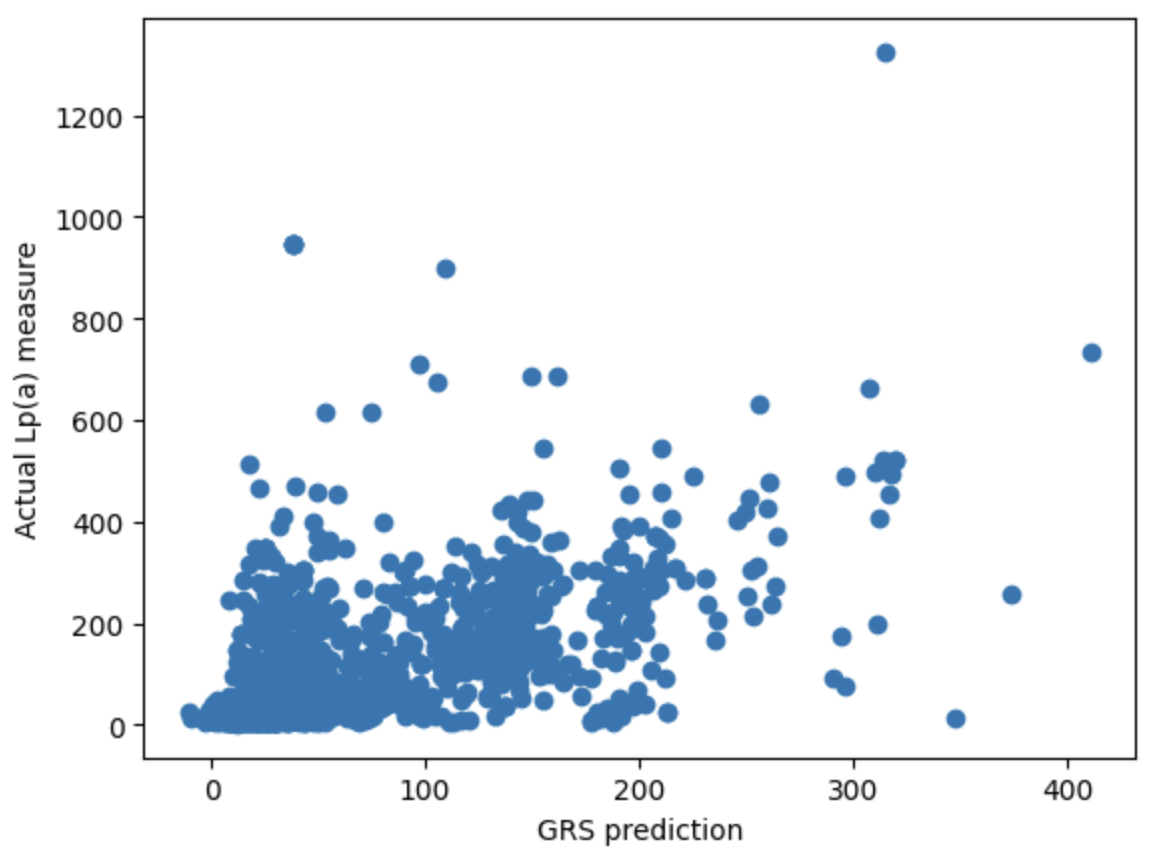

Figure 2. Scatterplot of KIV-2 normalized coverage & Lp(a) levels.

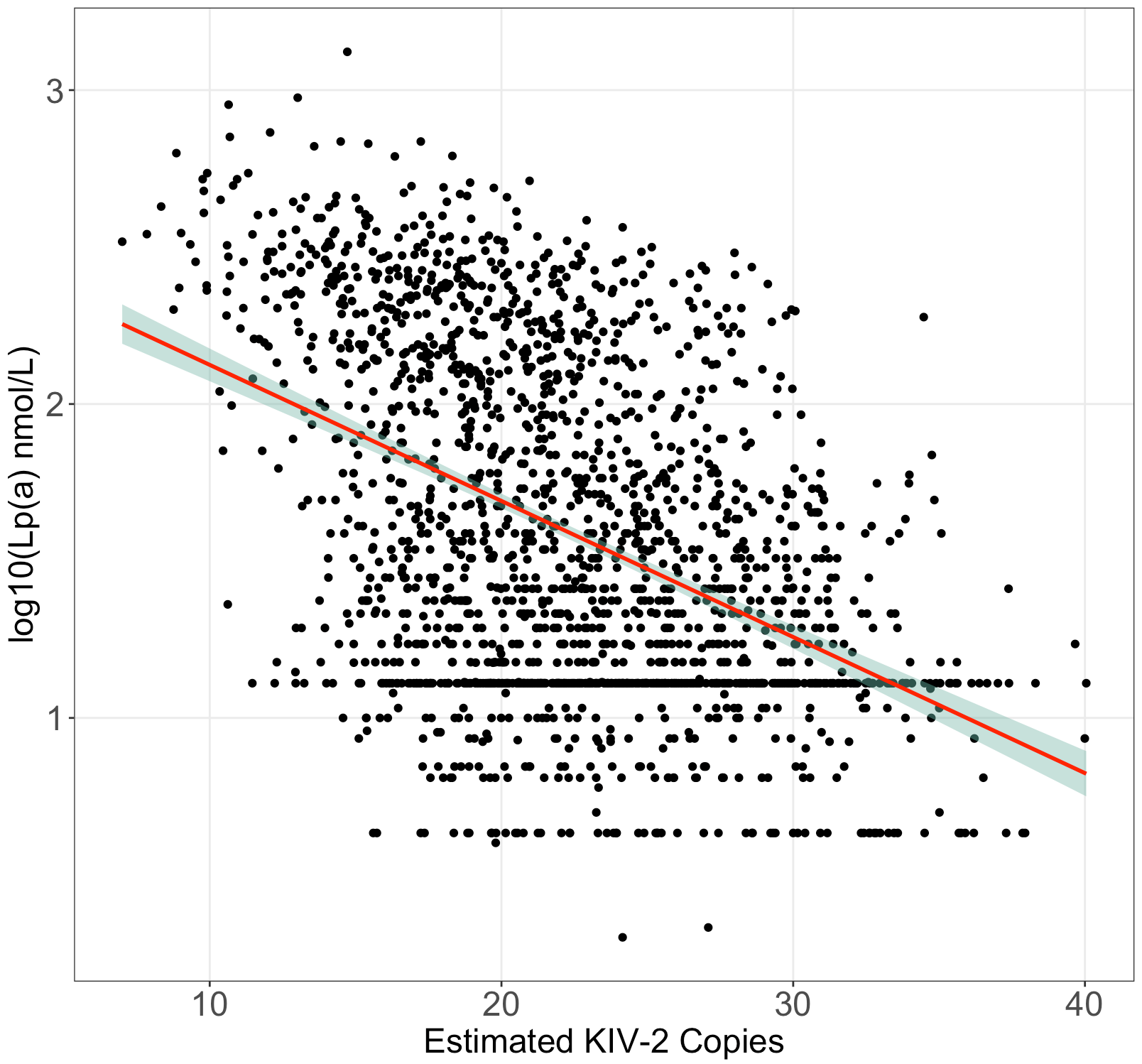

*p*=9.24×10^-96^, *R^2^*=20.16%

Figure 3. Distribution of KIV-2 normalized coverage by assay version.

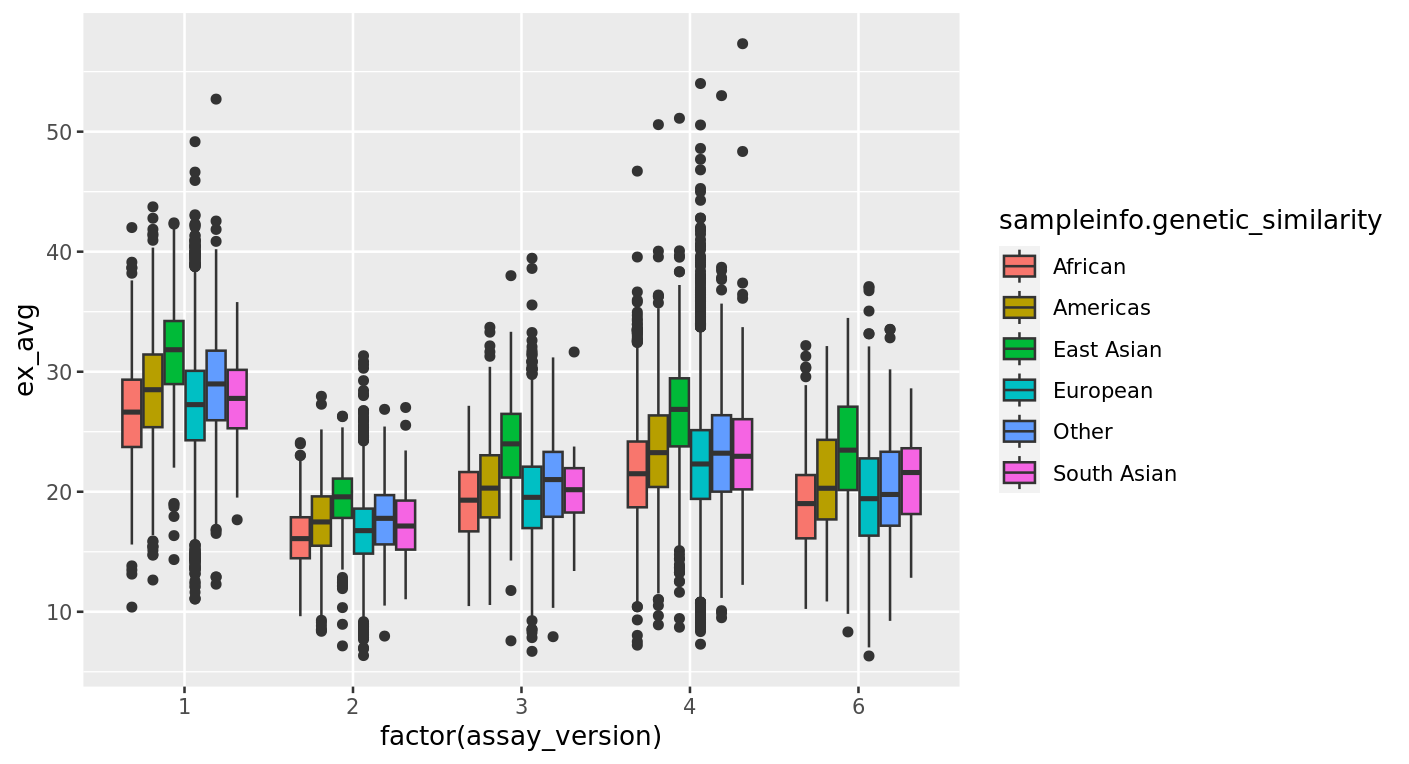

Figure 4. LDL measures and Lp(a) measures by genetic risk for a high Lp(a).

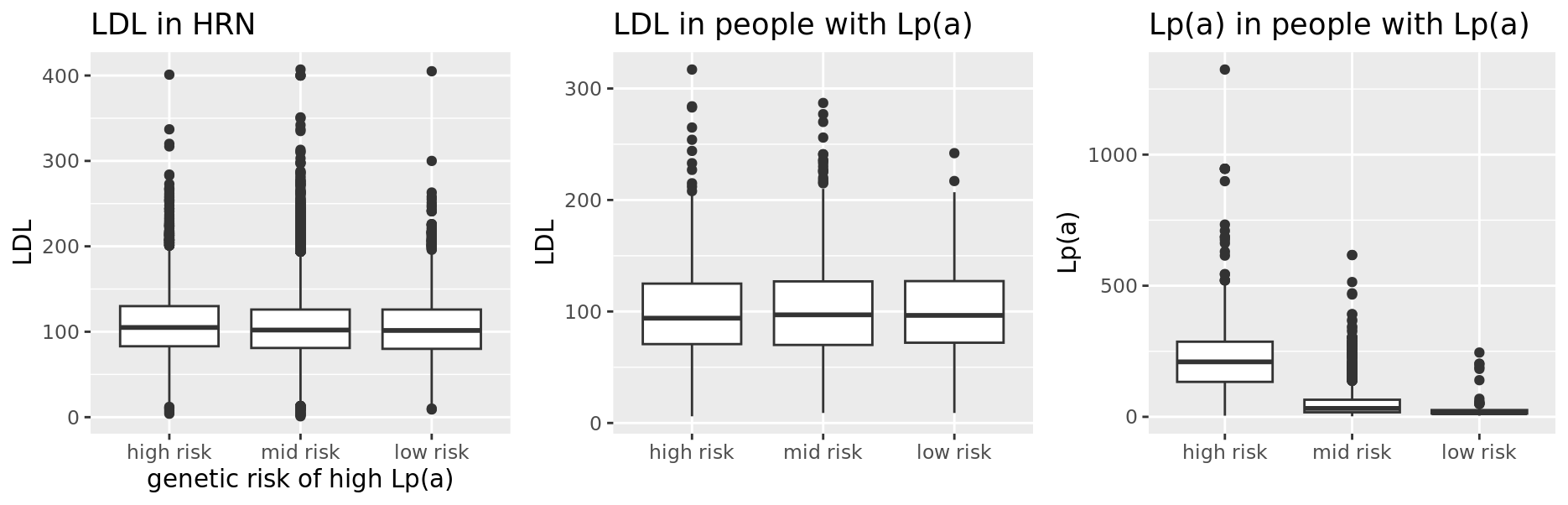

Figure 5. Average age in individuals by genetic risk for a high Lp(a).

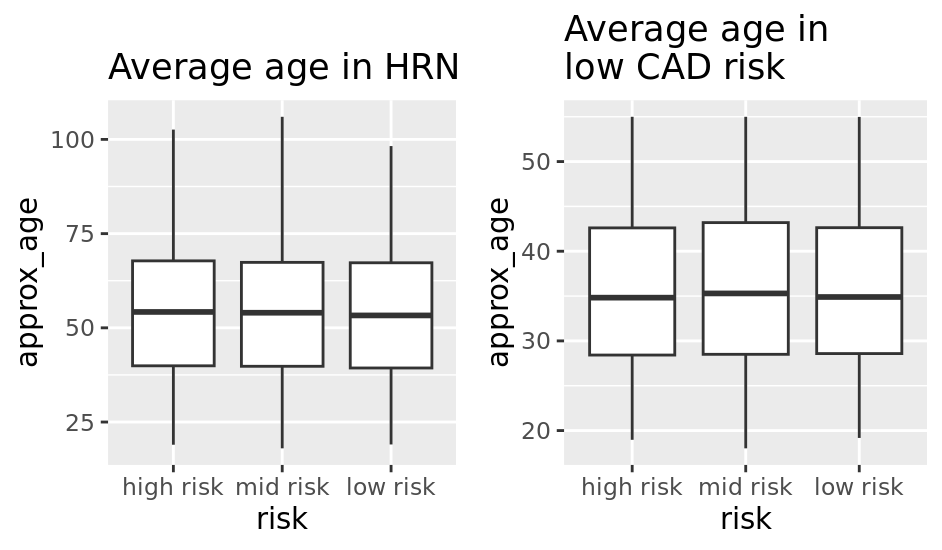

Table 1. Population demographics for all individuals with a calculated KIV-2 CNE.

| Self-reported sex | Pop: AFR | AMR | EAS | EUR | SAS | Other |
| --- | --- | --- | --- | --- | --- | --- |
| Male | 693 | 1993 | 553 | 18516 | 262 | 769 |
| Female | 2219 | 4990 | 1414 | 42890 | 246 | 1602 |

Table 2. Population demographics for all individuals with an Lp(a) measurement.

| Self-reported sex | Pop: AFR | AMR | EAS | EUR | SAS | Other |
| --- | --- | --- | --- | --- | --- | --- |
| Male | 15 | 56 | 12 | 550 | 10 | 15 |
| Female | 24 | 88 | 18 | 947 | 4 | 28 |

Table 3: Population counts for individuals in Lp(a) genetic risk brackets.

KIV-2 CNE low risk threshold >1.5SD

| Lp(a) genetic risk category | Pop: AFR | AMR | EAS | EUR | SAS | Other |
| --- | --- | --- | --- | --- | --- | --- |
| Average risk | 2373 | 3810 | 1323 | 43629 | 404 | 1594 |
| High risk | 338 | 2468 | 97 | 10916 | 56 | 480 |
| Low risk | 201 | 705 | 547 | 6861 | 48 | 297 |

KIV-2 CNE low risk threshold >2SD

| Lp(a) genetic risk category | Pop: AFR | AMR | EAS | EUR | SAS | Other |
| --- | --- | --- | --- | --- | --- | --- |
| Average risk | 2633 | 4449 | 1632 | 51892 | 453 | 1875 |
| High risk | 148 | 2024 | 77 | 4443 | 23 | 312 |
| Low risk | 131 | 510 | 258 | 5071 | 32 | 184 |

Appendix 1. OMOP ODHSI concept code IDs corresponding to measurement and phenotypic codesets.

Appendix 2. Cox proportional hazard estimates across phenotypes for high combined genetic risk of high Lp(a) vs average and low combined genetic risk of high Lp(a) in >120 nmol/L and >150nmol/L threshold groups.
